# Design and Validation of an AI-Assisted Sequential Screening Framework for Psychological Distress in Glaucoma

**DOI:** 10.64898/2026.05.20.26353679

**Authors:** Natalie A. Chou, Youngsoo Baek, Feilin Feng, Kaiwen Lu, Eun Young Choi, Hannah M. Fisher, Davina Malek, Alessandro Jammal, Tamara J. Somers, Kelly W. Muir, Felipe Medeiros, Samuel I. Berchuck

**Affiliations:** Department of Psychiatry and Behavioral Sciences, Duke University, Durham, North Carolina, USA; Department of Biostatistics & Bioinformatics, Duke University, Durham, North Carolina, USA; Department of Statistical Science, Duke University, Durham, North Carolina, USA; Kaiser Permanente, Santa Clara Homestead Medical Center, Santa Clara, California, USA; Bascom Palmer Eye Institute, University of Miami, Miami, Florida, USA; Massachusetts Eye and Ear, Harvard University, Boston, Massachusetts, USA; Duke Eye Center and Department of Ophthalmology, Duke University, Durham, North Carolina, USA

**Keywords:** Primary open-angle glaucoma, Psychological distress screening, Machine learning, Electronic health records, Clinical decision support

## Abstract

**Purpose:** Psychological distress is highly prevalent in glaucoma and is associated with worse adherence, reduced quality of life, and faster disease progression. However, distress is rarely assessed in ophthalmology settings due to time, workflow, and staffing constraints. We evaluated two artificial intelligence (AI)-based screening strategies, designed to efficiently identify distressed primary open angle glaucoma (POAG) patients during routine care, aiming to achieve effective, resource conscious, low burden clinical screening.

**Design:** Hybrid retrospective cohort and prospective cross-sectional study.

**Participants:** The retrospective cohort included >3,000 POAG patients from the Duke Ophthalmic Registry. Prospective validation was conducted in a separate 300 POAG patient cohort who completed patient-reported distress screening.

**Methods:** Using retrospective data, a neural network model was trained to predict an electronic health record (EHR)-derived computable phenotype of distress (“silver standard”). Prospective validation used the 8-item Patient Health Questionnaire (PHQ-8) as the “gold standard.” Three screening strategies were compared against PHQ-8: (1) universal PHQ-2 screening (two-item screener administered to all patients), (2) AI-only screening (fully automated EHR-based screener), and (3) sequential screening, (only patients flagged as high risk by AI screener completed the PHQ-2). Performance metrics included sensitivity, specificity, positive predictive value (PPV), negative predictive value (NPV), accuracy, and screening burden.

**Main Outcome Measures:** Sensitivity; specificity; PPV; NPV; accuracy; proportion of patients requiring secondary screening (screening burden).

**Results:** Distress prevalence was 17% (PHQ-8 > 6). Universal PHQ-2 screening (> 0) achieved high sensitivity (0.96) but lower specificity (0.73) and PPV (0.41), while requiring screening of all patients. The AI-assisted sequential approach substantially reduced screening burden while maintaining strong diagnostic performance. By administering PHQ-2 to ∼25% of patients, sequential screening achieved sensitivity 0.64, specificity 0.93, PPV 0.64, NPV 0.93, and accuracy 0.88, representing a ∼50% increase in PPV compared to PHQ-2 alone. AI-only screening reduced burden further but did not achieve comparable sensitivity or predictive performance.

**Conclusions:** AI-assisted sequential screening enables scalable, resource efficient identification of psychological distress in glaucoma care, substantially reducing screening burden while preserving clinically meaningful performance. This framework offers a practical pathway for integrating distress screening into routine ophthalmology workflows and improving the identification and referral of at-risk patients.

---

Primary open angle glaucoma (POAG) is a chronic, progressive optic neuropathy that affects over 65 million people aged 40-80 worldwide,^1^ and remains the leading cause of irreversible vision loss.^2^ As such, the uncertainty about future vision loss and the disease’s gradual progression contribute to considerable psychological burden in patients.^3^ While management primarily targets intraocular pressure (IOP) reduction to slow functional decline, factors such as emotional wellbeing, social support, and mental health, are increasingly recognized as critical in disease evaluation and care.^4,5^

The chronic course of POAG often leads to psychological distress including anxiety, depression, fear of blindness, and social isolation, all of which substantially reduce quality of life.^6–8^ Psychological distress prevalence in POAG patients is notably high, with up to 25% experiencing anxiety and 30% experiencing depression, rates more than 10% higher than the general population.^6^ Disease severity influences these outcomes, as worse functional disease severity^8^ and faster rates of progression^9^ correlate with greater depression. Importantly, psychological distress impacts disease management and prognosis. Patients reporting distress frequently show poor glaucoma comprehension,^10^ missed follow up visits,^11^ and suboptimal medication adherence,^12^ contributing to long term visual field loss. Psychological distress may additionally exert a direct physiologic effect, as stress can elevate IOP, the only modifiable risk factor in glaucoma.^4^ Experimental studies in primates^13^ and humans^14^ confirm that acute stress can increase IOP. Addressing psychological distress early in the treatment pathway could improve adherence, outcomes, and quality of life, but routine screening in glaucoma care is not common.

Early identification of psychological distress enables timely intervention and reduces adverse outcomes.^15^ However, conventional screening approaches are often limited by substantial time and cost demands.^16^ To address these challenges, several medical specialties have adopted more efficient strategies, including brief self-report questionnaires and two stage screening frameworks. In oncology and cardiology, for example, initial large scale pre-screening with short instruments, such as the National Comprehensive Cancer Network (NCCN) Distress Thermometer^17^ or the two item Patient Health Questionnaire^18^, is followed by more comprehensive assessment only in patients with positive results.^19^ This approach reduces resource burden while targeting high risk patients for further evaluation.^20^ In glaucoma care, however, routine psychological distress screening remains uncommon despite its potential to identify vulnerable patients and enable early intervention, representing a critical gap in supportive disease management. The most direct solution would be universal administration of validated patient reported outcome (PRO) measures. Our group has taken steps toward this goal, including recent work validating the Distress Thermometer in patients with POAG, supporting the feasibility and clinical relevance of routine distress screening in this population.^21^ Nevertheless, universal screening has not been widely implemented in ophthalmology practice due to persistent workflow, time, and resource constraints. As a result, there is a need for scalable approaches that approximate the benefits of universal screening while remaining feasible in routine care. In this context, we propose an AI-assisted screening framework designed to reduce screening burden while preserving performance comparable to universal screening.

Electronic health record (EHR) data have been widely used for predictive modeling across clinical domains,^22–25^ including work demonstrating accurate prediction of incident suicide risk.^26^ These advances highlight the potential of artificial intelligence (AI) to enable scalable, population level identification of patients at risk for adverse outcomes. In prior work, we developed machine learning models using EHR data from a large retrospective glaucoma cohort and demonstrated strong discrimination for a computable distress phenotype (area under the receiver operating characteristic [ROC] curve > 0.90).^27^ However, because patient reported distress measures were not available in that setting, outcomes were defined using an EHR-based proxy (hereafter referred to as a “silver standard”). While such approaches are useful for large scale model development, understanding clinical impact requires evaluation against validated PROs that directly measure psychological distress.

To address this gap, we conducted a prospective study with PRO measures of distress, enabling evaluation against a gold standard in a clinical setting. We evaluated whether AI-based screening strategies can approximate the performance of universal screening while reducing implementation burden. Specifically, we compared three approaches for identifying psychological distress: (1) universal screening using the two item Patient Health Questionnaire (PHQ-2), (2) fully automated AI-only screening based on EHR data, and (3) an AI-assisted sequential screening strategy in which the algorithm is used to pre-screen patients for targeted questionnaire administration. This sequential approach leverages AI to reduce screening burden by limiting manual questionnaire screening to a subgroup of high-risk patients, rather than relying on algorithmic predictions alone. This design enables direct evaluation of the tradeoff between screening burden and diagnostic performance across approaches. Together, these analyses provide a practical framework for implementing a scalable, clinically actionable distress screening in glaucoma care.

## METHODS

### Study Design

This study used data from both a large retrospective EHR cohort and a prospective cohort with PROs. The retrospective cohort was used to develop machine learning models using an EHR-derived phenotype of distress (silver standard), while the prospective cohort was used to evaluate model performance against PRO measures of distress (gold standard). Retrospective data were obtained from the Duke Ophthalmic Registry, which includes patients evaluated at the Duke Eye Center and affiliated satellite clinics between 2012 and 2021. Eligible patients were adults aged ≥18 years with POAG. Prospective data were obtained from a study examining patient attitudes toward psychological distress screening in glaucoma clinics.^21,28^ This study was approved by the Duke University Institutional Review Board (Pro00108155). The retrospective component was conducted under a waiver of informed consent due to the use of existing clinical data, and all participants in the prospective study provided informed consent. All procedures adhered to the tenets of the Declaration of Helsinki and complied with the Health Insurance Portability and Accountability Act.

**Figure 1** provides an overview of the study design, including development of the AI algorithm in the retrospective cohort and external validation in the prospective cohort. **Figure 2** summarizes the three screening strategies evaluated: (1) universal screening, (2) AI-only screening, and (3) AI-assisted sequential screening.

**Figure 1.**
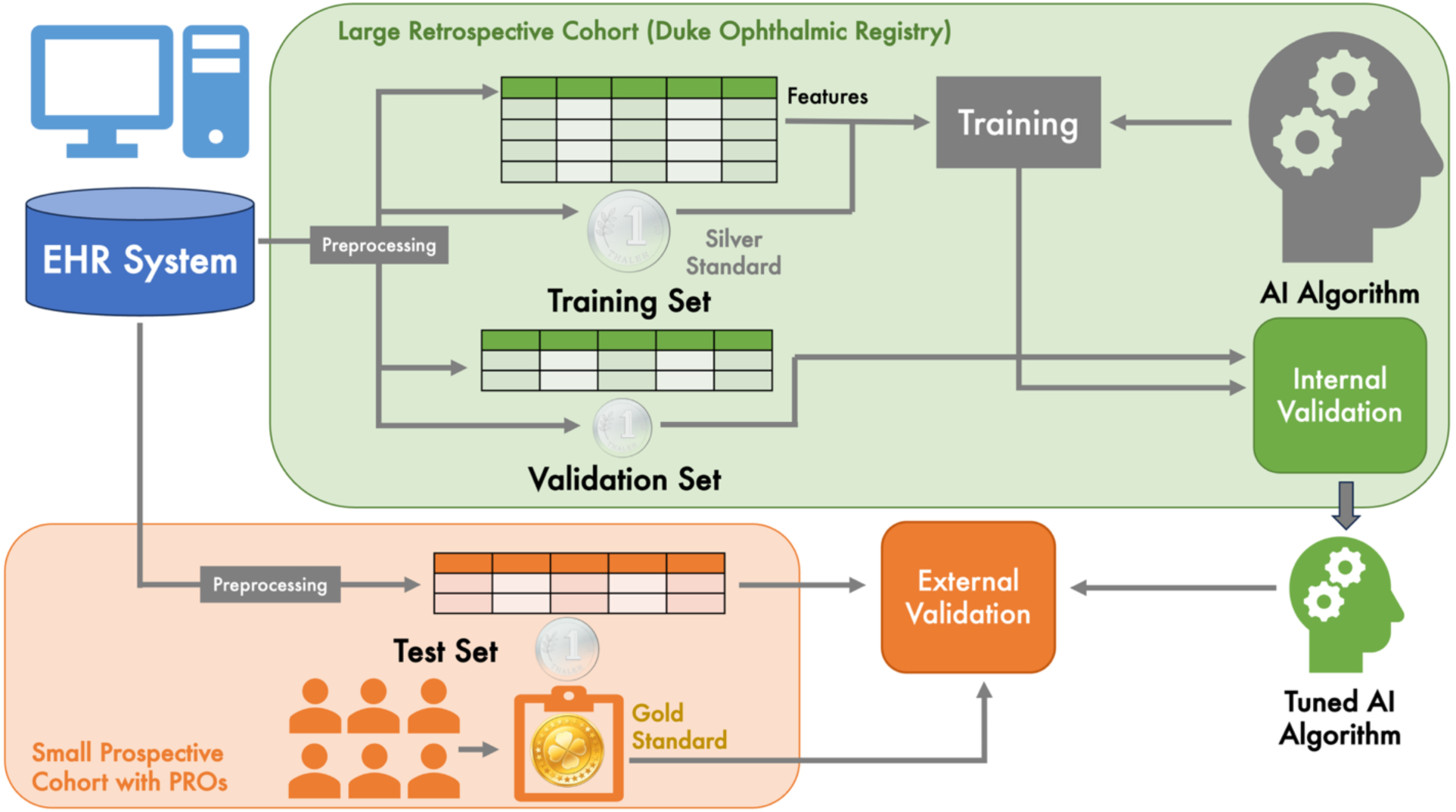
Schematic of Data Collection and Machine Learning Workflow. In the first phase, an AI model was trained on electronic health record (EHR) data from a retrospective cohort (Duke Ophthalmic Registry) to predict a silver standard outcome derived from EHR data. Model tuning and internal validation were performed within the retrospective dataset. In the second phase, the trained model was externally validated in a prospective cohort, based on its predictions for both silver standard outcomes and gold standard patient-reported outcomes (PROs).

**Figure 2.**
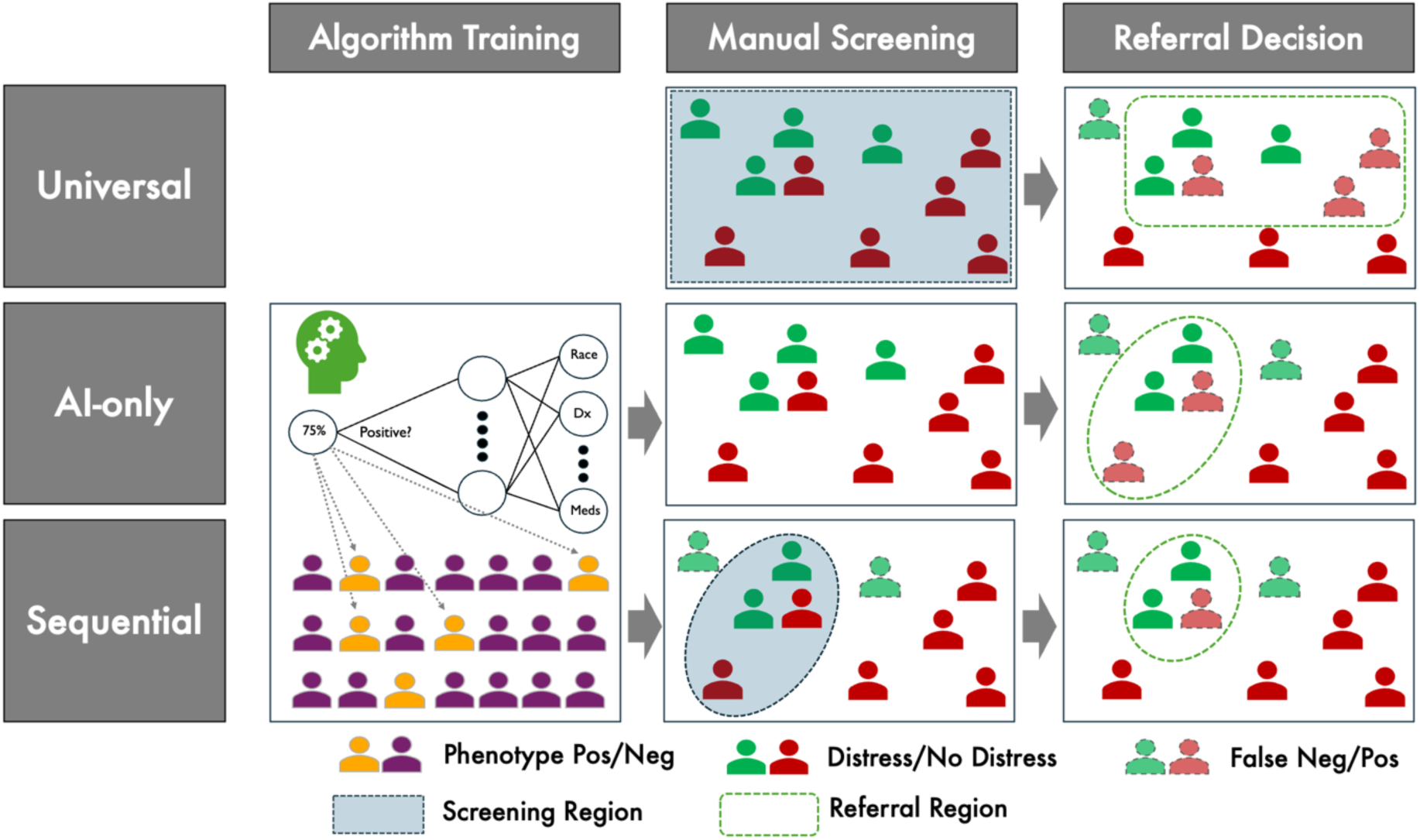
Comparison of Screening Strategies. Three approaches to identifying psychological distress are shown: (1) *Universal* screening, in which a patient-reported outcome (PRO) is administered to all patients; (2) *AI-only* screening, which relies exclusively on the trained AI model (Figure 1) to classify patients; and (3) *Sequential* screening, in which the AI model first identifies a subset of high-risk patients who then undergo targeted screening with a brief PRO. In Stage 1 (algorithm training), yellow and purple figures represent patients with positive and negative silver standard distress phenotypes. In Stage 2 (manual screening), green and red figures represent patients with and without distress based on the gold standard. Blue shading covers patients who receive PROs under each strategy. Green dashed line covers patients ultimately classified as having distress for referral.

### Retrospective Cohort and Model Development

The retrospective cohort was derived from the Duke Ophthalmic Registry and included adults (aged ≥18 years) with POAG who were evaluated at the Duke Eye Center and affiliated satellite clinics between 2012 and 2021. Patients were required to have at least two encounters and a minimum of one year of follow up. To align with the prospective validation cohort, analyses were restricted to encounters in the glaucoma clinic. POAG was defined using International Classification of Diseases diagnostic codes (H40.1), and the unit of analysis was the clinical encounter.

Psychological distress was defined using an encounter-level EHR-derived computable phenotype based on a validated Phenotype KnowledgeBase (PheKB) depression algorithm, as described in prior work.^27^ Because this definition relies on proxy clinical data rather than direct patient report, it is referred to as a silver standard measure of distress. Predictors were constructed from structured EHR data available prior to each encounter and included demographics, healthcare utilization (diagnoses, procedures, medications, and encounters), and problem-list information, resulting in a high-dimensional feature set for model development (1,844 variables). Socioeconomic variables, including income and education, were derived from U.S. Census data and linked at the Zip Code level. Income level was measured by per capita income in the past 12 months and was race specific. Education was measured by the percentage of residents who achieved a high school education and was sex specific. Insurance status (commercial, Medicaid, Medicare, uninsured, other) was also included.

Two predictive models were trained to estimate encounter-level probabilities of psychological distress: an elastic net classifier and a feedforward neural network. The elastic net model was fit using patient-level 10-fold nested cross-validation to select optimal regularization parameters and prevent overfitting. The neural network consisted of a four-layer architecture with ReLU activation functions and was trained using the Adam optimizer,^29^ with early stopping to improve generalization.^30^ To enhance stability, predictions were averaged across multiple random initializations. Model performance was evaluated using the area under the receiver operating characteristic (ROC) curve and precision-recall (PR) curve in held-out data constructed via patient-level splitting. These models generated the predicted probabilities used for AI-only screening and as the first stage of the sequential screening strategy.

### Prospective Cohort and Outcome Assessment

Patients with POAG were recruited from the Duke Eye Center glaucoma clinics between September 2022 and August 2023. Study procedures have been described previously and are briefly summarized here.^21,28^ Eligibility required age ≥18 years, a confirmed POAG diagnosis, at least one prior glaucoma clinic visit, and a minimum of one year of follow up. Exclusion criteria included recent glaucoma or cataract surgery within 6 months, reported or suspected serious mental illness (e.g., psychosis), or legal blindness, as determined by provider or chart review. Eligible patients were contacted by email prior to scheduled clinic visits, and consenting participants completed a set of questionnaires.

Demographic and clinical characteristics were collected to describe the cohort. Patient reported measures included age, sex, race, ethnicity, marital status, education, income, and employment status. Clinical data were obtained from the Duke Ophthalmic Registry and supplemented by manual chart review. Eye specific measures included intraocular pressure (IOP) and standard automated perimetry mean deviation (SAP MD), reported for the better eye.

Psychological distress was assessed using the Patient Health Questionnaire-8 (PHQ-8), which served as the gold standard measure of distress.^18^ The PHQ-8 includes all items from the PHQ-9 except for the question assessing suicidal or self-injurious thoughts; this item was excluded to avoid triggering risk assessments that could not be reliably and sufficiently managed in a remote research setting. Prior validation studies have demonstrated that the removal of this item has minimal impact on total scores and does not alter established cutoffs for depression severity.^31,32^ The PHQ-8 evaluates the frequency of depressive symptoms over the prior two weeks, with total scores ranging from 0 to 24. Consistent with prior work, clinically relevant psychological distress was defined as a PHQ-8 score > 6.^33,34^ The PHQ-8 was used for evaluation purposes and was not part of the screening strategies assessed in this study.

### Screening Strategies

We evaluated three approaches for identifying psychological distress: (1) universal questionnaire-based screening, (2) AI-only screening, and (3) an AI-assisted sequential screening strategy. Universal screening was conducted using the two item Patient Health Questionnaire (PHQ-2), a brief validated screener for depressive symptoms. In accordance with American Heart Association (AHA) recommendations for depression screening,^18^ any positive response (score > 0) was considered indicative of a positive screen. In the AI-only approach, our EHR-based algorithm was used to assign each patient a predicted probability of psychological distress based on routinely collected clinical data. Patients with predicted probabilities above a specified threshold were classified as distressed (more details in the following Statistical Evaluation section). This approach is fully automated and does not require administration of patient questionnaires, relying solely on EHR-derived predictions.

Finally, the AI-assisted sequential strategy combines algorithmic risk stratification with targeted questionnaire-based assessment. In this approach, the AI algorithm is first used to identify patients at elevated risk for psychological distress. Only these high-risk patients then undergo questionnaire-based screening using the PHQ-2, and patients with a positive PHQ-2 result are classified as distressed. This design follows established two stage screening frameworks used in other clinical domains, such as oncology and cardiology,^35,36^ while reducing the burden of universal screening by limiting questionnaire administration to a subset of high-risk patients.

### Statistical Evaluation

Summary statistics for cohort characteristics are presented using mean and standard deviation (SD) for continuous variables and counts and percentages for categorical variables. Hypothesis tests are presented, with categorical variables tested using a Chi-square test, and continuous variables tested using a Wilcoxon rank sum test. Model discrimination was evaluated using ROC and precision-recall (PR) curves in both the training (retrospective) and test (prospective) datasets. In the training data, performance was assessed against the EHR-derived silver standard. In the test data, performance was evaluated against both the silver standard and the patient reported gold standard. Areas under the curve were computed for both ROC and PR curves.

Screening strategies were evaluated using the prospective cohort with PROs as the gold standard (PHQ-8 > 6). Diagnostic performance was assessed using sensitivity, specificity, positive predictive value (PPV), negative predictive value (NPV), and overall accuracy. We considered two complementary evaluation approaches reflecting distinct clinical objectives. First, to assess whether AI-only screening could serve as a substitute for universal questionnaire-based screening, thresholds for the AI model were selected to match the specificity of PHQ-2 screening (score > 0). This enabled a direct comparison of sensitivity, predictive values, and accuracy under comparable operating conditions. Results are presented for both elastic net and neural network models. Second, for the AI-assisted sequential strategy, we aimed not to replicate the operating characteristics of universal screening, but to evaluate the tradeoff between screening burden and diagnostic performance. In this setting, thresholds for the AI model were selected using Youden’s index^37^ to balance sensitivity and specificity. The PHQ-2 threshold indicating distress was fixed at any score > 0. Screening burden was defined as the proportion of patients requiring questionnaire-based screening, which varies with the chosen AI threshold; operating characteristics were evaluated as a function of this proportion to characterize the tradeoff between burden and performance. Operating characteristics for the sequential strategy were further examined across demographic subgroups, including age, sex, and race.

Variable importance was computed for both models. For the elastic net model, the largest 25 coefficients in absolute value were presented along with their odds ratios (OR). OR p-values were not presented, since they cannot be computed reliably for the elastic net model.^38^ The non-zero demographic predictors were also presented. For a measure of variable importance in neural network, Shapley values^39^ were computed on a random subset of 1,000 encounters from the EHR data. Variables with the top 25 absolute Shapley values for positive prediction were presented, along with a list of demographic variables with the top 10 largest values.

All analyses were conducted using R (Version 4.0.5, R Core Team, Vienna, Austria) and Python (Version 3.13, Python Software Foundation) within the Protected Analytics Computing Environment (PACE) at Duke University. The glmnet and PyTorch packages were used for model development.^40–42^

## RESULTS

The retrospective cohort consisted of 31,043 encounters from 3,149 patients with a mean (SD) of 9.9 (7.3) encounters per patient over 2.6 (2.2) years of follow up. The mean age of the patients at first encounter was 68.6 (13.2) years with a breakdown of 1,790 (57%) females, 1,651 (52%) White, 1,337 (42%) Black/African American, and the rest Asian, multiracial, and other races. There were 1,240 (39%) patients with at least one encounter with a positive silver standard distress phenotype. The prospective cohort consisted of 300 patients with an average age of 68.4 (12.5) years, with a breakdown of 155 (52%) females, 242 (81%) White, and 46 (15%) Black/African American. Full summary details for the retrospective and prospective cohorts can be found in **Table 1** and **Table 2**, respectively.

**Table 1.** Patient-level summary statistics for the training set data derived from a retrospective cohort. Summaries are presented across the silver standard distress status (at least one encounter with a positive EHR-derived distress phenotype). P-values represent hypothesis tests across distress. Continuous variables were tested using Wilcoxon rank sum test and categorical variables with Chi-square exact test.

| Variable | All | Distress | No Distress | P-Values |
| --- | --- | --- | --- | --- |
| n | 3,149 | 1,240 (39%) | 1,909 (61%) |  |
| Number of encounters |  |  |  |  |
| Mean (SD) | 9.86 (7.32) | 9.92 (7.37) | 9.82 (7.28) |  |
| Median [min, max] | 8 [1, 46] | 8 [1, 41] | 8 [1, 46] |  |
| Follow-up in years, Mean (SD) | 4.1 (2.50) | 4.06 (2.49) | 4.13 (2.51) |  |
| Age (years) | 68.57 (13.17) | 69.08 (12.48) | 68.23 (13.59) | 0.075 |
| Race (%) |  |  |  | 0.002 |
| White | 1,651 (52%) | 698 (56%) | 953 (50%) |  |
| Black/African American | 1,337 (42%) | 487 (39%) | 850 (45%) |  |
| Other | 161 (5%) | 55 (4%) | 106 (6%) |  |
| Sex (Female, %) | 1,790 (57%) | 788 (64%) | 1,002 (52%) | <0.001 |
| Ethnicity (Hispanic/Latino, %) | 55 (2%) | 20 (2%) | 35 (2%) | 0.679 |
| Marital Status (Single, %) | 1,479 (47%) | 639 (52%) | 840 (44%) | <0.001 |
| Alcohol Use (%) | 1,568(50%) | 617(50%) | 951(50%) | 1.000 |
| Smoking Use (%) | 1,448(46%) | 639(52%) | 809(42%) | <0.001 |
| Illicit Drug Use (%) | 117(4%) | 73(6%) | 44(2%) | <0.001 |
| High School Education in %, Mean (SD) | 87 (0.13) | 87 (0.12) | 87 (0.13) | 0.826 |
| Annual Income in \$1k, Mean (SD) | 32.28 (17.32) | 32.47 (17.43) | 32.16 (17.25) | 0.628 |
| Insurance |  |  |  | <0.001 |
| Commercial (%) | 611 (19%) | 186 (15%) | 425 (22%) |  |
| Medicaid (%) | 97 (3%) | 47 (4%) | 50 (3%) |  |
| Medicare (%) | 2,399 (76%) | 995 (80%) | 1,404 (74%) |  |
| Uninsured (%) | 42 (1%) | 12 (1%) | 30 (2%) |  |

**Table 2.** Summary statistics for the test set data derived from a prospective cohort. Summaries are presented across gold standard distress status (PHQ-8 > 6). P-values represent hypothesis tests across distress. Continuous variables were tested using Wilcoxon rank sum test and categorical variables with Chi-square test.

| Variable | All | Distress | No Distress | P-Values |
| --- | --- | --- | --- | --- |
| n | 300 | 50 (17%) | 250 (83%) |  |
| PHQ-8, Mean (SD) | 3.51 (3.99) | 10.74 (3.89) | 2.07 (1.89) |  |
| PHQ-2 |  |  |  |  |
| Mean (SD) | 0.71 (1.15) | 2.52 (1.49) | 0.34 (0.61) |  |
| >0 (%) | 39 | 96 | 27 |  |
| Age (years) | 68.36 (12.45) | 62.7 (16) | 69.49 (11.32) | <0.001 |
| Race (%) |  |  |  | 0.064 |
| Caucasian/white | 242 (81%) | 35 (70%) | 207 (83%) |  |
| Black/African American | 46 (15%) | 13 (26%) | 33 (13%) |  |
| Other | 12 (4%) | 2 (4%) | 10 (4%) |  |
| Sex (Female, %) | 155 (52%) | 29 (58%) | 126 (50%) | 0.355 |
| Ethnicity (Hispanic/Latino, %) | 5 (2%) | 0 (0%) | 5 (2%) | 0.594 |
| Marital Status (Single, %) | 90 (30%) | 24 (48%) | 66 (26%) | 0.004 |
| Education (High school or lower, %) | 73 (24%) | 17 (34%) | 56 (22%) | 0.103 |
| Income (Less than 100k/year) |  |  |  | 0.010 |
| % | 139 (55%) | 31 (74%) | 108 (51%) |  |
| Missing | 48 (16%) | 8 (16%) | 40 (16%) |  |
| Employment (%) |  |  |  | 0.001 |
| Retired | 173 (58%) | 25 (50%) | 148 (59%) |  |
| Employed | 102 (34%) | 13 (26%) | 89 (36%) |  |
| Disability | 14 (5%) | 7 (14%) | 7 (3%) |  |
| Other | 11 (4%) | 5 (10%) | 6 (2%) |  |
| IOP (mm Hg) |  |  |  | 0.026 |
| Mean (SD) | 11.02 (3.59) | 10.31 (3.79) | 11.16 (3.54) |  |
| Missing | 2 (1%) | 0 (0%) | 2 (1%) |  |
| Visual Acuity (logMAR) |  |  |  | 0.962 |
| Mean (SD) | 0.18 (0.32) | 0.26 (0.45) | 0.16 (0.29) |  |
| Missing | 2 (1%) | 0 (0%) | 2 (1%) |  |
| SAP MD (dB) |  |  |  | 0.713 |
| Mean (SD) | -4.07 (5.86) | -4.63 (6.19) | -3.95 (5.79) |  |
| Missing | 29 (10%) | 6 (11%) | 23 (9%) |  |
| Canceled Appointment (%) |  |  |  | 0.166 |
| Not cancelled | 249 (83%) | 37 (74%) | 212 (85%) |  |
| Patient | 34 (11%) | 9 (18%) | 25 (10%) |  |
| Provider | 17 (6%) | 4 (8%) | 13 (5%) |  |

dB = Decibel; IOP = intraocular pressure; mm Hg = Milimeters mercury; logMAR = logarithm of minimum angle of resolution; PHQ-2 = patient health questionnaire 2-item scale; PHQ-8 = PHQ 8-item scale; SAP MD = Standard automated perimetry mean deviation; SD = Standard deviation

**Figure 3** presents ROC and PR curves for both machine learning models evaluated in the training (retrospective) and test (prospective) data, against both silver and gold standard outcomes. Both models demonstrated strong discrimination when predicting the silver standard, with ROC AUCs of 0.93 (neural network) and 0.86 (elastic net) in the training data, and 0.83 and 0.86, respectively, in the test data. PR AUCs were 0.89 and 0.79 in training, and 0.79 for both models in the test data. When evaluated against the gold standard in the test data, discrimination was lower, reflecting the increased difficulty of predicting PROs. However, the neural network retained meaningful predictive signal, with ROC AUC of 0.76 and PR AUC of 0.39, and outperformed the elastic net (ROC AUC 0.64; PR AUC 0.30). For comparison, universal PHQ-2 screening achieved higher discrimination (ROC AUC 0.94; PR AUC 0.80). Given the superior performance of the neural network for the gold standard outcome, subsequent analyses focus on the neural network model, with elastic net results reported for completeness.

**Figure 3.**
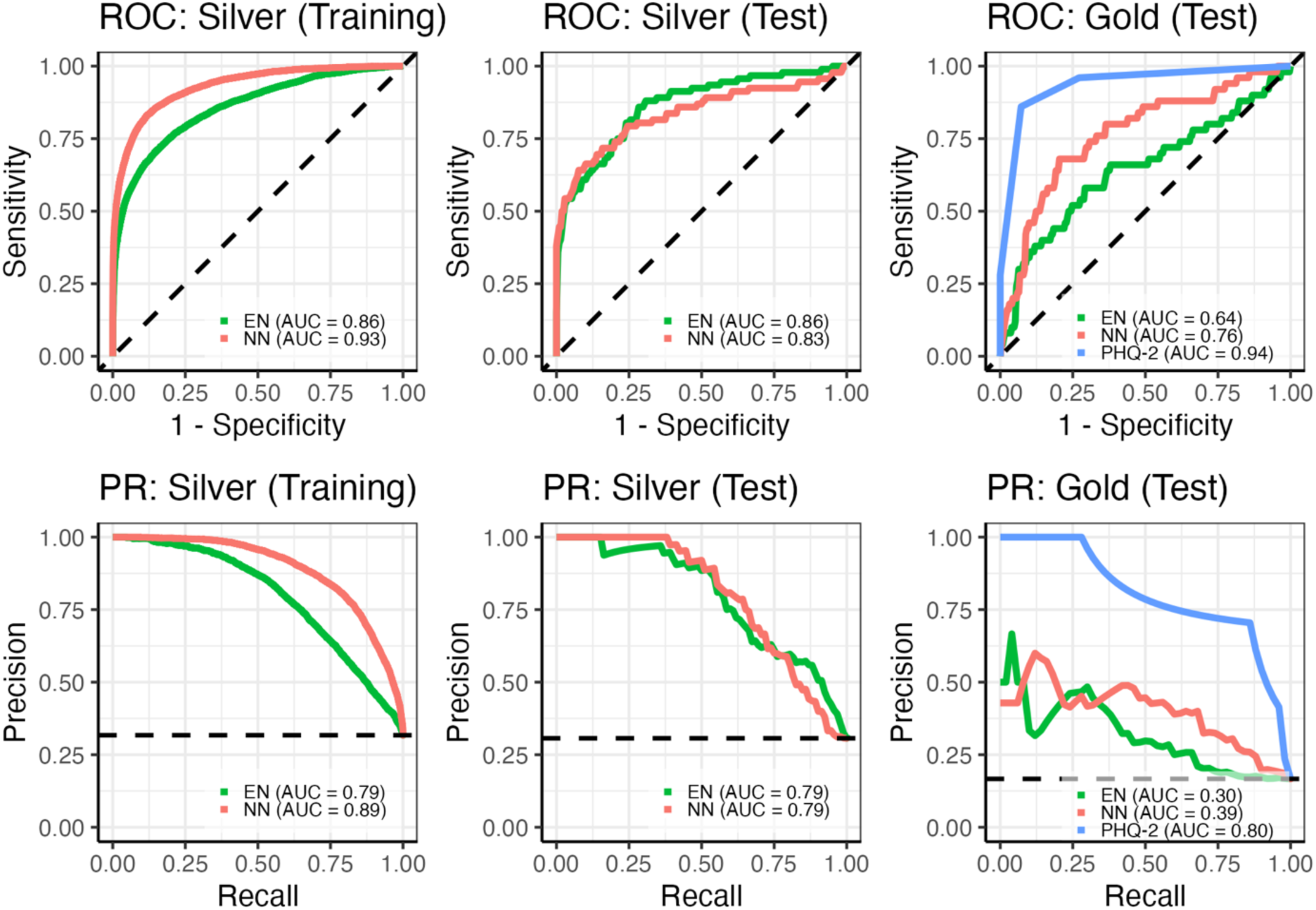
Receiver operating characteristic (ROC; top row) and precision-recall (PR; bottom row) curves for distress prediction models in training and test data. The left and middle columns show performance for predicting the silver standard distress phenotype in the training and test sets, respectively, comparing neural network (NN) and elastic net (EN) models. The right column shows performance for predicting gold-standard distress (PHQ-8 > 6) in the test set, comparing NN, EN, and PHQ-2 screening. Area under the curve (AUC) values are reported for each model and outcome.

**Table 3** compares the diagnostic performance of the three screening approaches. Universal screening using PHQ-2 achieved high sensitivity (0.96) and NPV (0.99), with specificity of 0.73, PPV of 0.41, and accuracy of 0.77, but required questionnaire administration for all patients. In contrast, AI-only screening eliminated the need for questionnaire-based screening but did not achieve comparable diagnostic performance. The neural network-based screening, with the threshold fixed to achieve the same specificity of 0.73 (anyone with a predicted probability of distress ≥ 21%), yielded sensitivity of 0.68, PPV of 0.33, NPV of 0.92, and accuracy of 0.72. The sequential screening strategy, using the Youden’s index-based threshold (anyone with a predicted probability of distress ≥ 26%) provided a balance between these approaches. When using the neural network, sequential screening achieved the highest specificity (0.93), PPV (0.64), and accuracy (0.88), while maintaining high NPV (0.93), with only a modest reduction in sensitivity (0.64). This improvement was achieved while limiting questionnaire-based screening to 28% of patients, compared to 100% under universal screening. **Table 4** presents the diagnostic characteristics across subgroups of age, sex and race for the sequential screening method relying on the neural network. The same breakdown is given for the elastic net model in **Supplementary Table S1**. These findings highlight a tradeoff between sensitivity and operational feasibility, with sequential screening achieving improved precision while substantially reducing screening burden.

**Table 3.** Operating characteristics of manual, AI-only, and sequential screening strategies for identifying elevated distress (PHQ-8 > 6). Manual screening was defined as PHQ-2 > 0. AI-only strategies (neural network / elastic net) were evaluated under two different thresholding methods, applied to each predicted probability of distress: (1) thresholds selected to match the specificity of manual screening (specificity = 0.73) to enable direct comparison, and (2) thresholds selected using Youden’s index to optimize discrimination. Sequential screening used the AI model (threshold selected by Youden’s index) as a first-stage triage tool, with PHQ-2 administered only to patients identified as high risk. Proportion requiring screening denotes the fraction of participants receiving PHQ-2.

| Screening Approach | Specificity | Sensitivity | PPV | NPV | Accuracy | Proportion<br>Requiring<br>Screening | Threshold |
| --- | --- | --- | --- | --- | --- | --- | --- |
| *Manual* |  |  |  |  |  |  |  |
| PHQ-2 | 0.73 | 0.96 | 0.41 | 0.99 | 0.77 | 1.00 | Score > 0 |
| *AI-only (specificity matched to PHQ-2)* |  |  |  |  |  |  |  |
| Neural Network | 0.73 | 0.68 | 0.33 | 0.92 | 0.72 | 0.00 | Probability<br>≥ 21% |
| Elastic Net | 0.73 | 0.52 | 0.28 | 0.88 | 0.69 | 0.00 | Probability<br>≥ 29% |
| *AI-only (threshold based on Youden's index)* |  |  |  |  |  |  |  |
| Neural Network | 0.80 | 0.68 | 0.40 | 0.93 | 0.78 | 0.00 | Probability<br>≥ 26% |
| Elastic Net | 0.71 | 0.58 | 0.28 | 0.89 | 0.69 | 0.00 | Probability<br>≥ 28% |
| *Sequential* |  |  |  |  |  |  |  |
| Neural Network | 0.93 | 0.64 | 0.64 | 0.93 | 0.88 | 0.28 | Probability<br>≥ 26% |
| Elastic Net | 0.90 | 0.54 | 0.52 | 0.91 | 0.84 | 0.34 | Probability<br>≥ 28% |

NPV = Negative Predicted Value; PHQ-2 = Patient Health Questionnaire 2-item scale; PPV = Positive Predicted Value

**Table 4.** Operating characteristics of the neural network sequential screening strategy by demographic subgroup. Counts, number of patients with distress, and distress prevalence are also reported for each subgroup.

| Variable | n | Distress | Specificity | Sensitivity | PPV | NPV | Accuracy |
| --- | --- | --- | --- | --- | --- | --- | --- |
| Gender |  |  |  |  |  |  |  |
| Male | 145 | 21 (14%) | 93 | 48 | 53 | 91 | 86 |
| Female | 155 | 29 (19%) | 93 | 76 | 71 | 94 | 90 |
| Race |  |  |  |  |  |  |  |
| White | 242 | 35 (14%) | 91 | 71 | 58 | 95 | 88 |
| Black/African American | 46 | 13 (28%) | 100 | 46 | 100 | 83 | 85 |
| Other | 12 | 2 (17%) | 100 | 50 | 100 | 91 | 92 |
| Age (years) |  |  |  |  |  |  |  |
| <50 | 23 | 9 (39%) | 93 | 67 | 86 | 81 | 83 |
| 50-60 | 33 | 6 (18%) | 96 | 40 | 67 | 88 | 86 |
| 60-70 | 97 | 19 (20%) | 94 | 75 | 75 | 94 | 90 |
| 70-80 | 130 | 15 (12%) | 92 | 57 | 47 | 94 | 88 |
| >80 | 37 | 5 (14%) | 91 | 60 | 50 | 94 | 86 |

NPV = Negative Predicted Value; PPV = Positive Predicted Value.

**Figure 4** illustrates the tradeoff between screening burden and diagnostic performance for the neural network sequential strategy. The vertical dashed line indicates the operating point selected using Youden’s index, which corresponds to the results in **Table 3**. As the operating threshold varies, the proportion of patients requiring questionnaire-based screening changes, with corresponding changes in sensitivity, specificity, and predictive values. These curves demonstrate that the operating point can be adjusted to achieve different balances between screening burden and diagnostic performance. **Supplementary Figure S1** shows the same curves for elastic net.

**Figure 4.**
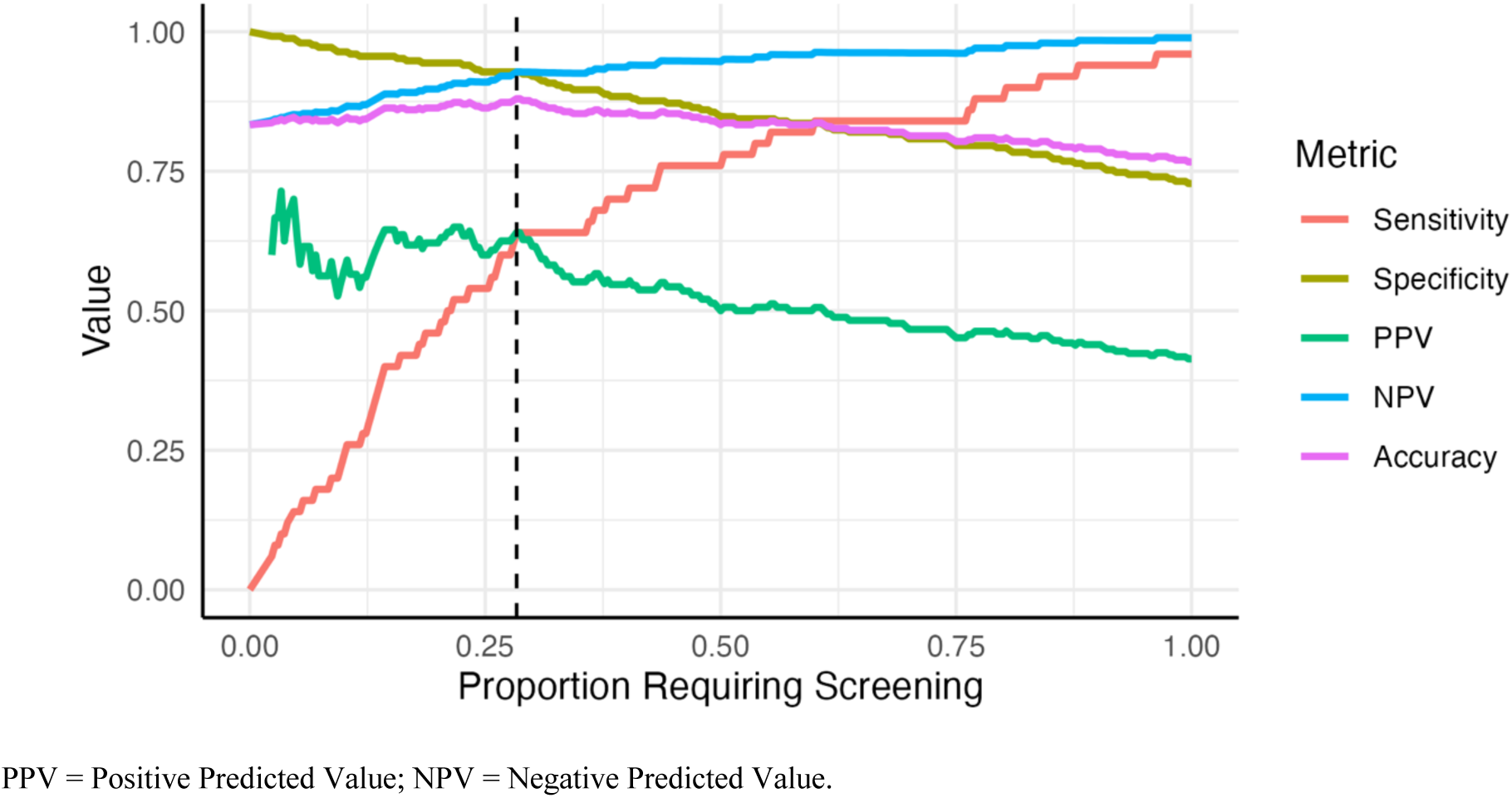
Operating characteristics of the neural network sequential screening strategy as a function of the proportion requiring manual screening. The vertical dashed line indicates the threshold corresponding to Youden’s index, which was used to compute the metrics in **Tables 3** and **4**.

Variable importance measures for each model are reported in the **Supplementary Tables S2-S4. Table S2** presents a list of variables with the 25 highest absolute Shapley values for the neural network predicting positive psychological distress; **Supplementary Table S3** contains the top 10 demographic predictors for the same model. Finally, **Supplementary Table S4** shows the top 25 predictors from the elastic net model ordered by the largest coefficients in absolute value.

## DISCUSSION

This study developed and validated AI-based screening strategies for identifying psychological distress in glaucoma care, including both a fully automated AI-only approach and an AI-assisted sequential screening framework. Our findings demonstrate that while AI-only screening does not achieve performance comparable to universal PHQ-2 administration, it captures meaningful predictive signal that can be leveraged to improve efficiency when combined with targeted PRO screening. The sequential approach improved specificity, PPV, and overall accuracy, while reducing the need for questionnaire-based screening by approximately fourfold, supporting a practical balance between diagnostic performance and real world feasibility.

A central challenge of implementing psychological distress screening in ophthalmology is balancing its diagnostic performance with the practical constraints of clinical workflows. Universal PHQ-2 screening maximizes sensitivity but requires screening all patients, which may be difficult to sustain in busy clinics with limited staff and competing priorities. Conversely, fully automated approaches reduce burden, but rely solely on EHR-derived information and may not capture patient reported distress with sufficient accuracy. These constraints motivate approaches that can selectively target screening efforts while preserving meaningful clinical performance.

Consistent with this framing, the AI-only approach demonstrated uniformly lower performance than universal PHQ-2 screening across sensitivity, PPV, NPV, and accuracy when evaluated at matched specificity. In contrast, the sequential framework improved PPV and overall accuracy while substantially reducing screening burden. Although universal screening achieved the highest sensitivity, it required manual questionnaire administration for all patients. AI-only screening eliminated this burden, but at the cost of reduced sensitivity and predictive performance. The neural network-based sequential approach, borrowing strengths of both strategies, achieved the highest specificity and accuracy, and markedly increased PPV to 0.64, compared to 0.41 for PHQ-2 alone (approximately a 50% relative increase). Subgroup analyses (**Table 4**) further show that PPV remained generally high across demographic groups, indicating that the improved precision of the sequential approach was consistent across populations.

A key advantage of the sequential screening framework is its higher PPV and specificity, ensuring that patients flagged by the method are more likely to benefit from downstream psychological resources. While both universal and sequential approaches aim at identifying subsets of patients for referral, the sequential framework reduces the number of patients who require initial questionnaire-based screening, focusing clinical attention earlier in the workflow on a smaller, higher-risk group. The sequential approach therefore yields a more targeted cohort for referral (e.g., to a social worker), optimizing the limited availability of mental health service providers and increasing the likelihood of meaningful intervention. Efficient allocation is particularly critical in under resourced settings, as it promotes equity by prioritizing those with greater need, reduces stigma associated with false positives, and strengthens provider confidence in referrals. Moreover, this framework can be readily adapted to other chronic diseases characterized by similar workflow and psychological care challenges^43^ and aligns with value based care principles, emphasizing efficient, personalized screening that balances diagnostic precision with real world feasibility. Finally, any AI model that outputs predicted probabilities of psychological distress can be seamlessly integrated into this framework, enabling further improvements in performance as models evolve.

Reduced sensitivity in the sequential approach reflects an intentional and pragmatic tradeoff to improve feasibility. As illustrated in **Figure 4**, thresholds that maximize sensitivity come at the cost of substantially lower specificity and PPV, whereas modestly higher thresholds yield meaningful gains in predictive value and screening efficiency. In this context, higher PPV is particularly valuable, as patients identified by the sequential method are more likely to have true psychological distress, increasing clinical confidence and the likelihood of effective follow up. Furthermore, higher specificity resulting from algorithmic preselection implies that most patients not at risk avoid unnecessary and potentially distressing assessment, preserving scarce mental health resources.

These tradeoffs are especially relevant in ophthalmology, where universal PRO-based screening is often impractical due to workflow constraints, limited staff capacity, and the absence of standardized referral pathways.^44^ In such settings, high sensitivity approaches, while ideal in theory, may become “assessment without action,” where positive screens do not reliably lead to follow up care.^45^ The sequential framework instead aligns with real world clinical practice by focusing screening and downstream care on a smaller subset of high-risk patients, increasing the likelihood that identified patients receive meaningful intervention. Evidence from glaucoma patients indicates strong interest in both distress screening and referral for psychological support, underscoring the importance of designing workflows that translate screening into actionable care.^28^

Identifying psychological distress is only the first step; effective implementation requires clear pathways for follow up. In academic and large health systems, referral to embedded social workers or integrated behavioral health providers represents the ideal pathway. However, in many ophthalmology practices, particularly private or resource limited settings, alternative approaches are needed. Physicians may recommend peer support resources or condition specific support groups for vision loss, such as those offered through the Glaucoma Research Foundation or local vision rehabilitation programs. Coordination with the patient’s primary care provider provides another accessible pathway, enabling further evaluation and referral within an established care relationship. Brief educational materials addressing distress, coping strategies, and available community resources can also be provided at the point of care. As noted in prior work, integrating mental health screening and referral into ophthalmology, even in resource limited settings, can meaningfully improve patient wellbeing and engagement in care.^46^ Future implementation studies should evaluate the effectiveness of these pathways across diverse practice settings.

Several limitations warrant consideration. First, the sequential approach reduces sensitivity, and some patients with psychological distress may be missed under resource constrained screening, highlighting the need for careful calibration of thresholds as clinical workflows evolve. Second, the prospective validation cohort was predominantly White, which may limit generalizability to more diverse populations, so subgroup differences in performance warrant further study in larger and more representative cohorts. Third, although external validation was conducted using prospective data, model development and validation were performed within a single health system, and evaluation in multisite settings is needed to establish broader generalizability. Finally, reliance on an EHR-derived phenotype in the training phase may not fully capture patient reported distress and may introduce misclassification, underscoring the importance of incorporating additional gold standard PRO data in future work. Continued evaluation in prospective implementation studies will be important to assess workflow integration, patient and provider acceptability, and the impact of screening on downstream clinical care.

In conclusion, this study presents a practical, resource aware framework for screening psychological distress in glaucoma care using EHR-derived AI. While AI-only screening does not replace universal PRO-based screening, an AI-assisted sequential screening strategy provides a meaningful balance between diagnostic performance and feasibility. By substantially reducing burden while improving PPV and specificity, this approach offers a realistic and scalable pathway for integrating psychological risk assessment into routine ophthalmology care and may be adaptable to other chronic diseases with similar workflow and behavioral health challenges.

## Data Availability

All consented data produced in the present study are available upon reasonable request to the authors. Unconsented EHR data is cannot be made available, since it is not ours to share.

---

**Supplementary Table S1.** Operating characteristics of the elastic net sequential screening strategy by demographic subgroup. Counts, number of patients with distress, and distress prevalence are also reported for each subgroup.

| Variable | n | Distress | Specificity | Sensitivity | PPV | NPV | Accuracy |
| --- | --- | --- | --- | --- | --- | --- | --- |
| Gender |  |  |  |  |  |  |  |
| Male | 145 | 21 (14%) | 93 | 29 | 40 | 88 | 83 |
| Female | 155 | 29 (19%) | 87 | 72 | 57 | 93 | 85 |
| Race |  |  |  |  |  |  |  |
| White | 242 | 35 (14%) | 89 | 57 | 47 | 92 | 84 |
| Black/African American | 46 | 13 (28%) | 94 | 46 | 75 | 82 | 80 |
| Other | 12 | 2 (17%) | 100 | 50 | 100 | 91 | 92 |
| Age (years) |  |  |  |  |  |  |  |
| <50 | 23 | 9 (39%) | 93 | 44 | 80 | 72 | 74 |
| 50-60 | 33 | 6 (18%) | 91 | 40 | 50 | 88 | 82 |
| 60-70 | 97 | 19 (20%) | 91 | 56 | 60 | 90 | 84 |
| 70-80 | 130 | 15 (12%) | 90 | 43 | 35 | 92 | 85 |
| >80 | 37 | 5 (14%) | 88 | 100 | 56 | 100 | 89 |

NPV = Negative Predicted Value; PPV = Positive Predicted Value.

**Supplementary Figure S1.**
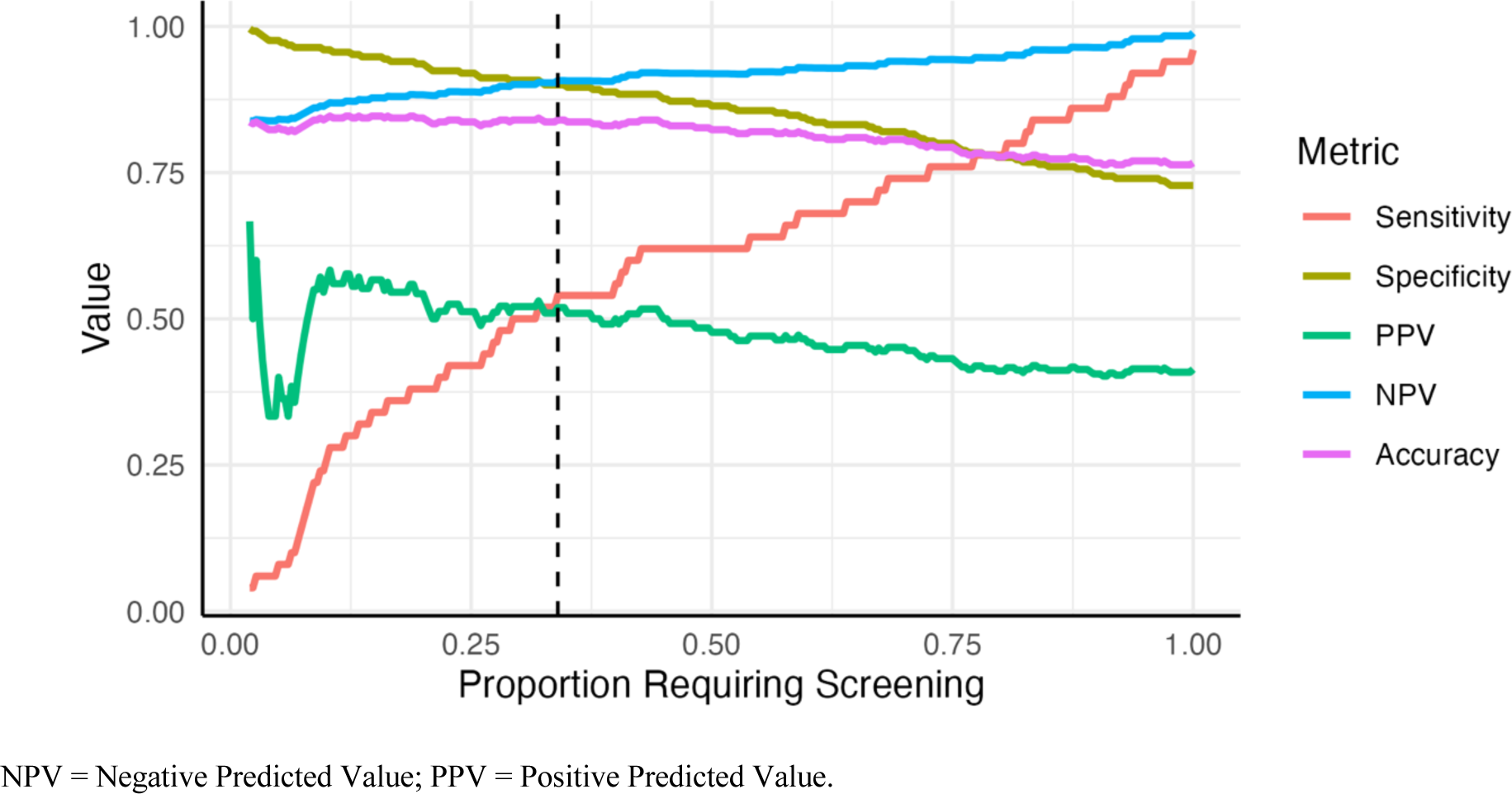
Operating characteristics of the elastic net sequential screening strategy as a function of the proportion requiring manual screening. The vertical dashed line indicates the threshold corresponding to Youden’s index, which was used to compute the metrics in **Tables 3** and **S1**.

**Supplementary Table S2.**
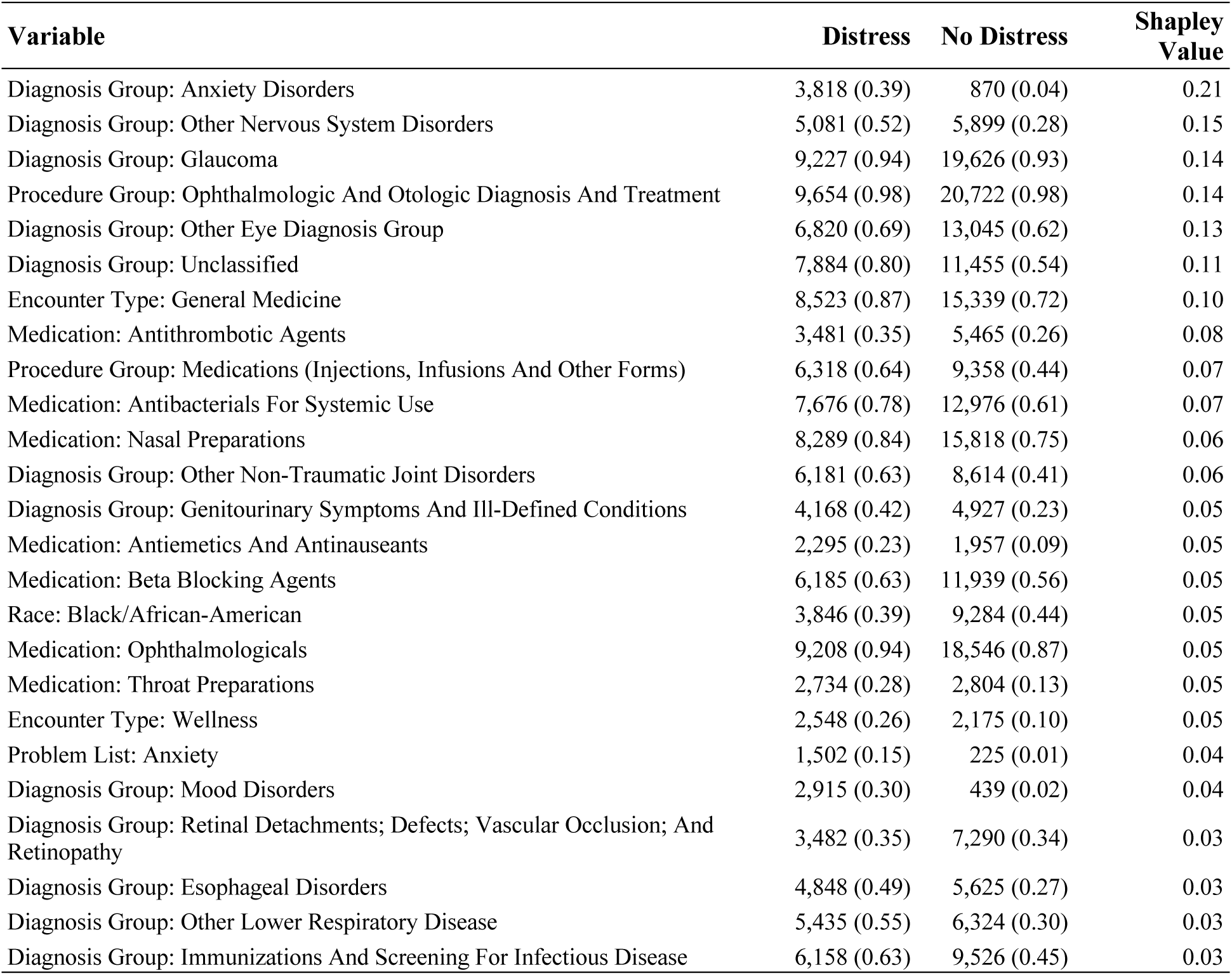
Predictors with the largest absolute Shapley values for positive prediction. All utilization variables presented used a 5-year window prior to the encounter. Also presented are the number of encounters with each predictor present, for both encounters with and without distress, along with corresponding proportions.

| Variable | Distress | No Distress | Shapley Value |
| --- | --- | --- | --- |
| Diagnosis Group: Anxiety Disorders | 3,818 (0.39) | 870 (0.04) | 0.21 |
| Diagnosis Group: Other Nervous System Disorders | 5,081 (0.52) | 5,899 (0.28) | 0.15 |
| Diagnosis Group: Glaucoma | 9,227 (0.94) | 19,626 (0.93) | 0.14 |
| Procedure Group: Ophthalmologic And Otologic Diagnosis And Treatment | 9,654 (0.98) | 20,722 (0.98) | 0.14 |
| Diagnosis Group: Other Eye Diagnosis Group | 6,820 (0.69) | 13,045 (0.62) | 0.13 |
| Diagnosis Group: Unclassified | 7,884 (0.80) | 11,455 (0.54) | 0.11 |
| Encounter Type: General Medicine | 8,523 (0.87) | 15,339 (0.72) | 0.10 |
| Medication: Antithrombotic Agents | 3,481 (0.35) | 5,465 (0.26) | 0.08 |
| Procedure Group: Medications (Injections, Infusions And Other Forms) | 6,318 (0.64) | 9,358 (0.44) | 0.07 |
| Medication: Antibacterials For Systemic Use | 7,676 (0.78) | 12,976 (0.61) | 0.07 |
| Medication: Nasal Preparations | 8,289 (0.84) | 15,818 (0.75) | 0.06 |
| Diagnosis Group: Other Non-Traumatic Joint Disorders | 6,181 (0.63) | 8,614 (0.41) | 0.06 |
| Diagnosis Group: Genitourinary Symptoms And Ill-Defined Conditions | 4,168 (0.42) | 4,927 (0.23) | 0.05 |
| Medication: Antiemetics And Antinauseants | 2,295 (0.23) | 1,957 (0.09) | 0.05 |
| Medication: Beta Blocking Agents | 6,185 (0.63) | 11,939 (0.56) | 0.05 |
| Race: Black/African-American | 3,846 (0.39) | 9,284 (0.44) | 0.05 |
| Medication: Ophthalmologicals | 9,208 (0.94) | 18,546 (0.87) | 0.05 |
| Medication: Throat Preparations | 2,734 (0.28) | 2,804 (0.13) | 0.05 |
| Encounter Type: Wellness | 2,548 (0.26) | 2,175 (0.10) | 0.05 |
| Problem List: Anxiety | 1,502 (0.15) | 225 (0.01) | 0.04 |
| Diagnosis Group: Mood Disorders | 2,915 (0.30) | 439 (0.02) | 0.04 |
| Diagnosis Group: Retinal Detachments; Defects; Vascular Occlusion; And Retinopathy | 3,482 (0.35) | 7,290 (0.34) | 0.03 |
| Diagnosis Group: Esophageal Disorders | 4,848 (0.49) | 5,625 (0.27) | 0.03 |
| Diagnosis Group: Other Lower Respiratory Disease | 5,435 (0.55) | 6,324 (0.30) | 0.03 |
| Diagnosis Group: Immunizations And Screening For Infectious Disease | 6,158 (0.63) | 9,526 (0.45) | 0.03 |

**Supplementary Table S3.**
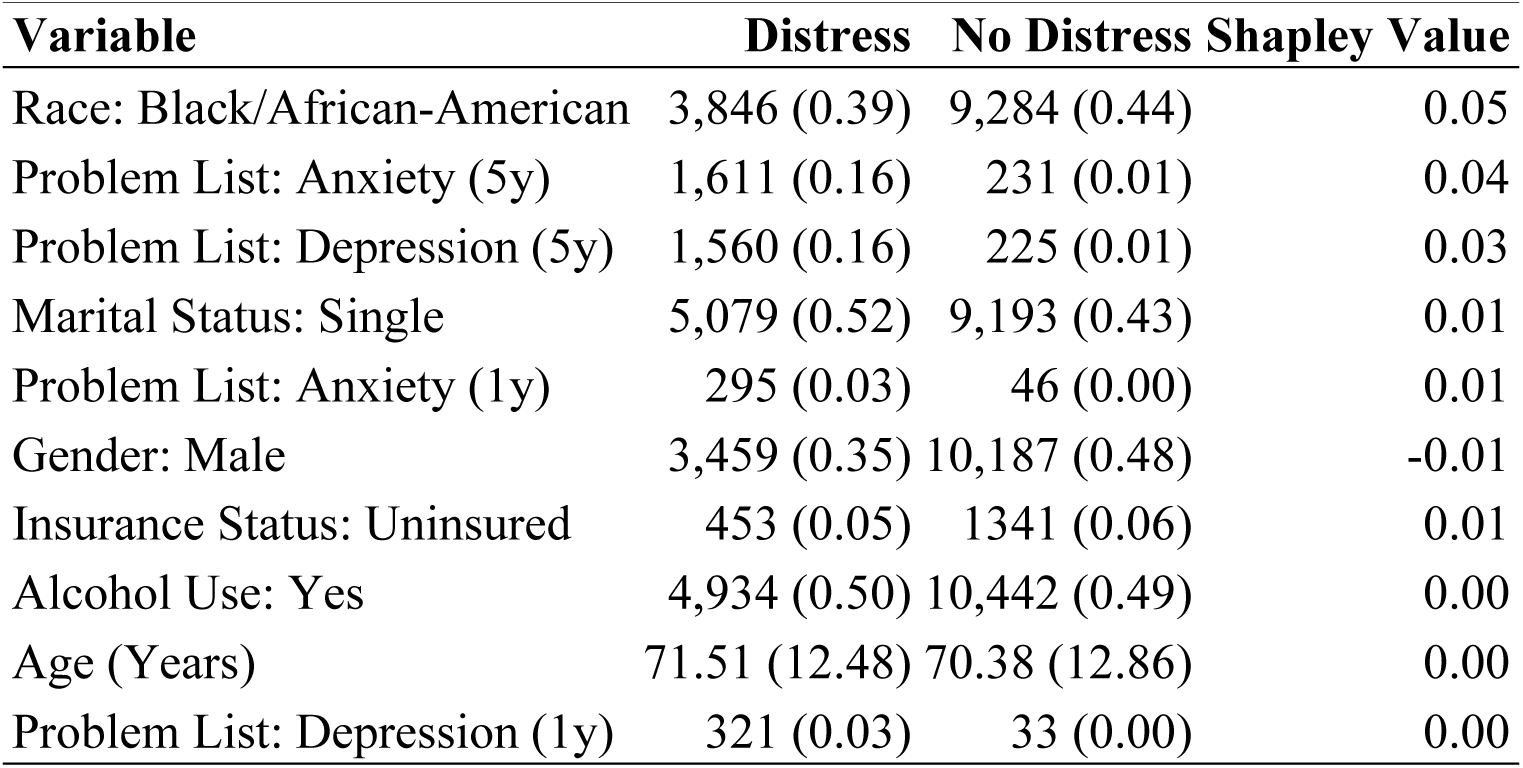
Demographic predictors with the largest absolute Shapley values for positive prediction.

**Supplementary Table S4.** Odds ratios (ORs) for the top 25 predictors of distress for elastic net, ordered by coefficient absolute values. In parenthases is the time window prior the encounter that the variable corresponds to, one year (1y) or five years (5y).

| Variable | Distress | No Distress | OR |
| --- | --- | --- | --- |
| Problem List: Depression (5y) | 1,497 (0.15) | 225 (0.01) | 4.15 |
| Problem List: Anxiety (5y) | 1,502 (0.15) | 225 (0.01) | 2.93 |
| Diagnosis Group: Anxiety disorders (5y) | 3,818 (0.39) | 870 (0.04) | 1.23 |
| Gender: Male | 3,459 (0.35) | 10,187 (0.48) | 0.85 |
| Drug Use: Yes | 588 (0.06) | 526 (0.02) | 1.16 |
| Smoking: Yes | 5,034 (0.51) | 8,817 (0.42) | 1.13 |
| Diagnosis Group: Mood Disorders (5y) | 2,915 (0.30) | 439 (0.02) | 1.11 |
| Diagnosis Group: Chest Pain (5y) | 3,896 (0.40) | 4,030 (0.19) | 1.03 |
| Encounter Type: Emergency Medicine (1y) | 3,200 (0.33) | 2,948 (0.14) | 1.03 |
| Medication: Psychoanaleptics (5y) | 4,797 (0.49) | 1,584 (0.07) | 1.03 |
| Procedure Group: Other non-OR therapeutic procedures, male genital (5y) | 76 (0.01) | 19 (0.00) | 1.03 |
| Diagnosis Group: Chronic obstructive pulmonary disease and bronchiectasis (1y) | 620 (0.06) | 363 (0.02) | 1.02 |
| Encounter Type: General Medicine (3m) | 5,062 (0.51) | 7,027 (0.33) | 1.02 |
| Diagnosis Group: Unclassified (1y) | 5,084 (0.52) | 6,062 (0.29) | 1.02 |
| Diagnosis Group: Substance-related disorders (5y) | 647 (0.07) | 192 (0.01) | 1.02 |
| Encounter Type: Wellness (5y) | 2,548 (0.26) | 2,175 (0.10) | 1.02 |
| Encounter Type: Radiology (5y) | 6,966 (0.71) | 10,841 (0.51) | 1.02 |
| Encounter Type: Infectious Diseases (5y) | 425 (0.04) | 358 (0.02) | 1.01 |
| Procedure Group: Spinal fusion (5y) | 570 (0.06) | 365 (0.02) | 1.01 |
| Procedure Group: Esophageal dilatation (5y) | 198 (0.02) | 57 (0.00) | 1.01 |
| Procedure Group: Cancer chemotherapy (3m) | 6,450 (0.66) | 9,878 (0.47) | 1.01 |
| Procedure Group: Other OR therapeutic procedures on skin and breast (5y) | 2,691 (0.27) | 2,304 (0.11) | 1.01 |
| Encounter Type: Infectious Diseases (1y) | 208 (0.02) | 101 (0.00) | 1.01 |
| Diagnosis Group: Mood disorders (1y) | 1,282 (0.13) | 130 (0.01) | 1.01 |
| Diagnosis Group: Fluid and electrolyte disorders (5y) | 1,301 (0.13) | 828 (0.04) | 1.01 |

